# Role of Multi-resolution Vulnerability Indices in COVID-19 spread: A Case Study in India

**DOI:** 10.1101/2021.07.19.21260791

**Authors:** Rupam Bhattacharyya, Anik Burman, Kalpana Singh, Sayantan Banerjee, Subha Maity, Arnab Auddy, Sarit Kumar Rout, Supriya Lahoti, Rajmohan Panda, Veerabhadran Baladandayuthapani

**Affiliations:** Department of Biostatistics, University of Michigan, Ann Arbor, USA; Indian Statistical Institute, Kolkata, India; Public Health Foundation of India, New Delhi, India; Operations Management & Quantitative Techniques Area, Indian Institute of Management, Indore, India; Department of Statistics, University of Michigan, Ann Arbor, USA; Department of Statistics, Columbia University, New York, USA; Indian Institute of Public Health, Bhubaneswar, Public Health Foundation of India, India

**Keywords:** COVID-19 Vulnerability, Vulnerability Indices, Health Indicators, Time-varying Reproduction Number, Bayesian Model Averaging

## Abstract

**Introduction:** The outbreak of COVID-19 has differentially affected countries in the world, with health infrastructure and other related vulnerability indicators playing a role in determining the extent of the COVID-19 spread. Vulnerability of a geographical region/country to COVID-19 has been a topic of interest, particularly in low- and middle-income countries like India to assess the multi-factorial impact of COVID-19 on the incidence, prevalence or mortality data.

**Datasets and Methods:** Based on publicly reported socio-economic, demographic, health-based and epidemiological data from national surveys in India, we compute contextual, COVID-19 Vulnerability Indices (cVIs) across multiple thematic resolutions for different geographical and spatial administrative regions. These multi-resolution cVIs were used in regression models to assess their impact on indicators of the spread of COVID-19 such as the average time-varying instantaneous reproduction number.

**Results:** Our observational study was focused on 30 districts of the eastern Indian state of Odisha. It is an agrarian state, prone to natural disasters and one of the largest contributors of an unprotected migrant workforce. Our analyses identified housing and hygiene conditions, availability of health care and COVID preparedness as important spatial indicators.

**Conclusion:** Odisha has demonstrated success in containing the COVID-19 infection to a reasonable level with proactive measures to contain the spread of the virus during the first wave. However, with the onset of the second wave of COVID, the virus has been making inroads into the hinterlands and peripheral districts of the state, burdening the already deficient public health system in these areas. The vulnerability index presented in this paper identified vulnerable districts in Odisha. While some of them may not have a large number of COVID-19 cases at a given point of time, they could experience repercussions of the pandemic. Improved understanding of the factors driving COVID-19 vulnerability will help policy makers prioritise resources and regions leading to more effective mitigation strategies for the COVID-19 pandemic and beyond.

WHAT IS ALREADY KNOWN
Measuring vulnerability to COVID-19 and other pandemics is a complex and layered subject. In Low-to-Middle-Income Country (LMIC) like India, complete reliance on incidence, prevalence or mortality data of the disease may not be the best measure since this data from the health system and DHS in public domain is limited.

ADDED VALUE OF THIS STUDY
To our knowledge, this is the first study at the district level concerning the COVID-19 situation in Odisha, characterized by a large tribal and migrant population. We defined vulnerability through relevant socio-economic domains that have an influence on mitigation strategies. Although we applied our methods to the districts of Odisha, we believe they can be used in other LMIC regions.

IMPLICATIONS OF THE FINDINGS
Regions with higher overall or theme-specific vulnerability index might experience potentially severe consequences of the COVID-19 outbreak demanding precise, dynamic and nimble policy decisions to prevent a potentially dire situation.

## INTRODUCTION

The outbreak of the highly infectious coronavirus disease 2019 (COVID-19), caused by the severe acute respiratory syndrome novel coronavirus (SARS-CoV-2), emerged in China and spread widely globally.^1^ As of July 16, 2021 India recorded more than 31 million confirmed cases, of which around 425,000 (1.4%) were active, more than 30 million (97.3%) recovered, and more than 412,000 (1.3%) have died.^2^ India, with a population of more than 1.34 billion, has a high population density of 454 people/km^2^, almost 3 times that of China,^3^ and faces multiple challenges to tackle COVID-19.^4^

While a lot of research has focused on clinical outcomes, epidemiological modelling, and transmission dynamics of the novel coronavirus,^5 6^ less focus has been placed on preponderance of risk and vulnerability to contracting the disease ascribed to social economic environment. Emerging studies have begun to report on the impacts of social vulnerability on COVID-19 from an incidence and outcome standpoint.^7 8^ However, the resolution of most studies has been at the global or country level, and less attention has been paid to a sub-regional or sub-national level. This is important in the Indian context as the *district* is the unit for administrative management and delivery of health services.^9^

Vulnerability in the present context of COVID-19 is more than just the risk of contracting the disease.^10^ It is described as a dynamic concept—a person or a group might not be vulnerable at the onset of the pandemic, but could subsequently become vulnerable depending on policies and response at the country or state level.^11^ The term vulnerability implies a measure of risk or consequences associated with socio-economic factors and financial, mental, and physical coping mechanisms resulting from a system’s ability to cope with the pandemic.^5 12^ Vulnerability manifests itself in different forms. Beyond epidemiological vulnerability to COVID-19 (e.g. elderly people and individuals with comorbidities), low and middle income countries (LMICs) such as India are characterized by other concerns - transmission vulnerability (e.g. population density, household and social structures, mobility, livelihood imperatives, hygiene infrastructure), health system vulnerability (e.g. availability of formal health care providers, intensive care), and vulnerability to direct control measures (e.g. impact of quarantines, lockdowns, self-isolation, disrupted social interactions).^13^ Poor public health infrastructure places India at a strategic disadvantage in this pandemic since it increases the risk of exposure to vulnerability.^14 15^

To this end, we created an interactive algorithm to estimate the COVID-19 Vulnerability Indices (cVI’s) across multiple geographical districts to COVID-19 and inform our understanding of the most potentially vulnerable district(s). We hypothesize that more vulnerable districts are less able and prepared to prevent, detect, and respond to COVID-19 spread, whereas more-resilient districts are better able to do so. We computed vulnerability indices across five factors - socioeconomic and demographic composition, housing and hygiene conditions, availability of health-care facilities, preparedness of COVID-19, and epidemiological factors.^10 16^ These factors were then individually and collectively assessed to identify geographical locations that are disproportionally exposed to the risk of infection and/or disease severity over the pandemic timeline across the districts of Odisha an eastern state in India which has reasonably managed to contain the virus in spite of poor infrastructure and limited resources.

## DATASETS AND METHODS

### Data description

District level data on the social, demographic, economic, housing & hygiene, health, preparedness of COVID-19, and epidemiological profile for the state of Odisha were collected from national and state-level surveys and dashboards in India, as summarized in Supplementary Table 1.^17–23^ We expanded the concept and computed vulnerability under different thematic domains, extending beyond social vulnerability to accommodate vulnerability contextual to the COVID-19 pandemic. More specifically, we considered five themes, which are important in the context of the COVID-19 epidemic in India, particularly for management of COVID-19 infection in the community, as described in Figure 1 and Supplementary Table 1. The datasets were available in different formats and at different spatial resolution, necessitating pre-processing and/or modelling before being input to the framework for constructing the cVI’s.

**Figure 1:**
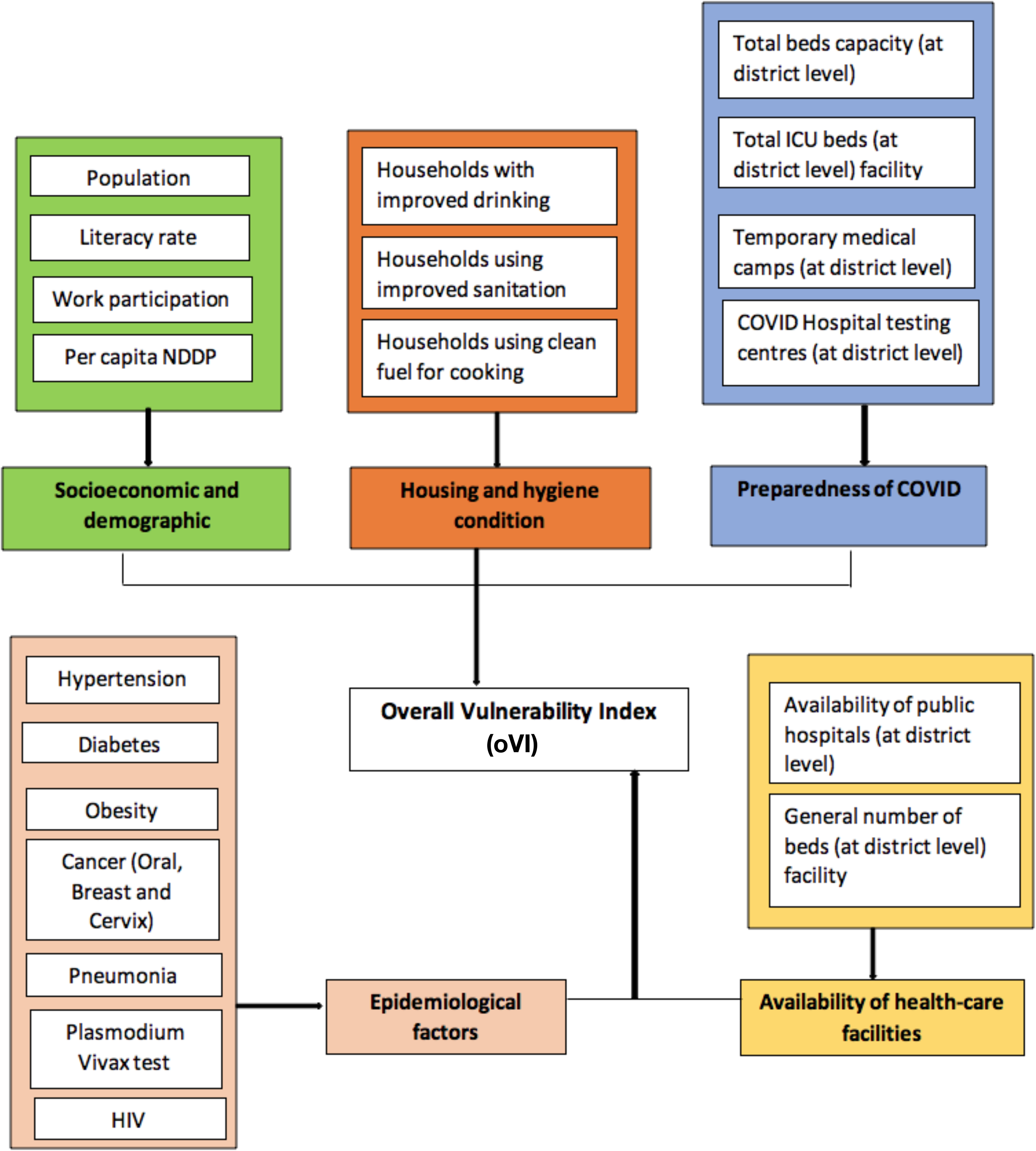
Summary of the five themes and the underlying variables used to compute the vulnerability indices. Each primary rectangle connected to the central overall vulnerability index box contains the name of one theme, and the corresponding sub-labels list the indicators included within that theme. All data sources are summarized in Supplementary Table 1.

The COVID-19 epidemic might be associated with far reaching and prolonged socioeconomic and demographic consequences. Epidemic-induced economic shock will mostly be felt by the middle and lower-income sections of society,^24^ and therefore it is important to consider the socioeconomic and demographic condition of a population when creating a vulnerability index. We used indicators relating to population dynamics since it affects the rate of transmission of the virus,^25^ education since it affects cognitive functioning, understanding of health and utilisation of healthcare,^26^ and also status of work due to its influence on resources and possession of assets as a proxy for poverty. In total, four indicators (Figure 1 and Supplementary Table 1) were used to define the Socioeconomic demographics. Furthermore, Housing and hygiene conditions, particularly uncontaminated and safe drinking water sources, healthy sanitation behaviour, and clean fuel for cooking, are important factors in COVID-19 transmission since they form the first line of defence against the infection.^27^ Exposure to smoke associated with the use of unclean or solid fuels is associated with harmful health effects as well and thus constitutes an important domain of vulnerability.^19^ We considered a total of three indicators in this group, namely, clean drinking water, non-shared toilet facilities and clean fuel for cooking (Figure 1 and Supplementary Table 1).

Two indicators were used to define availability of healthcare (Figure 1 and Supplementary Table 1). The management of an epidemic and the treatment seeking ability of a population depend on easy and affordable access to well capacitated health-care systems and health security, and thus should be included in the vulnerability index. The preparedness of COVID-19 was represented by four indicators (Figure 1 and Supplementary Table 1). The ability of a country/region to respond to the pandemic depends heavily on the healthcare systems capacity and preparedness.^28^ Besides, there are several known epidemiological factors and underlying medical conditions that might put a population at risk of higher morbidity and mortality from COVID-19 infection and thus merit inclusion in the vulnerability index. We considered six such indicators to define epidemiological factors (Figure 1 and Supplementary Table 1).

District-level COVID-19 count data on confirmed cases, deaths and recoveries were obtained from the COVID-19 India Dashboard.^2^

### Computation of relative COVID-19 vulnerability indices

The district-level covariates considered for the analyses varied in their scales and interpretations and covered a range of continuous (e.g., per capita Net District Domestic Product i.e., NDDP), count (e.g., district-wise population) and percentage/proportion (e.g., percentage of households with improved sanitation) data. The cVI’s were computed following a similar methodology to that developed by Flanagan and colleagues, used by the Center for Disease Control to compute social vulnerability indices for each census track in the USA.^29^ For the purposes of downstream interpretations and analyses, we aggregated these variables into unified vulnerability indices offering two major advantages. First, transforming them to a common scale of values between 0 to 1, with lower values (closer to 0) indicative of low vulnerability, and analogously higher values (closer to 1) indicative of higher vulnerability allows coherent interpretations across the different indices. Second, the common scale also allows us to extract regression coefficients and other associated metrics at comparable scales from models with more stable fits, as we show below.

We considered a total of 25 district-level indicators grouped into five broad themes, as described in the previous section and in Figure 1 and Supplementary Table 1. The amount of risk associated with a particular value of an indicator is interpreted as how detrimental its effects are on the population. The risk is higher towards the terminal value of an indicator where it is potentially causing a worse influence. In other words, the risk is high for high values of those indicators (covariates) which have negative influence on the population (e.g., disease proportion for districts); likewise, the risk is low for high values of those indicators which have a positive influence (e.g., hospital/bed counts). We adapt the algorithm by Acharya and Porwal, 2020 for the calculation of the relative vulnerability indices for each district.^10^ The algorithm consists of three steps, as summarized in the left panel of Figure 2 and mathematical derivations are provided in Supplementary Text Equations 1-3. We compute *indicator-specific relative cVI*, *theme-specific cVI* and an *overall vulnerability index (oVI)* for each district.

**Figure 2:**
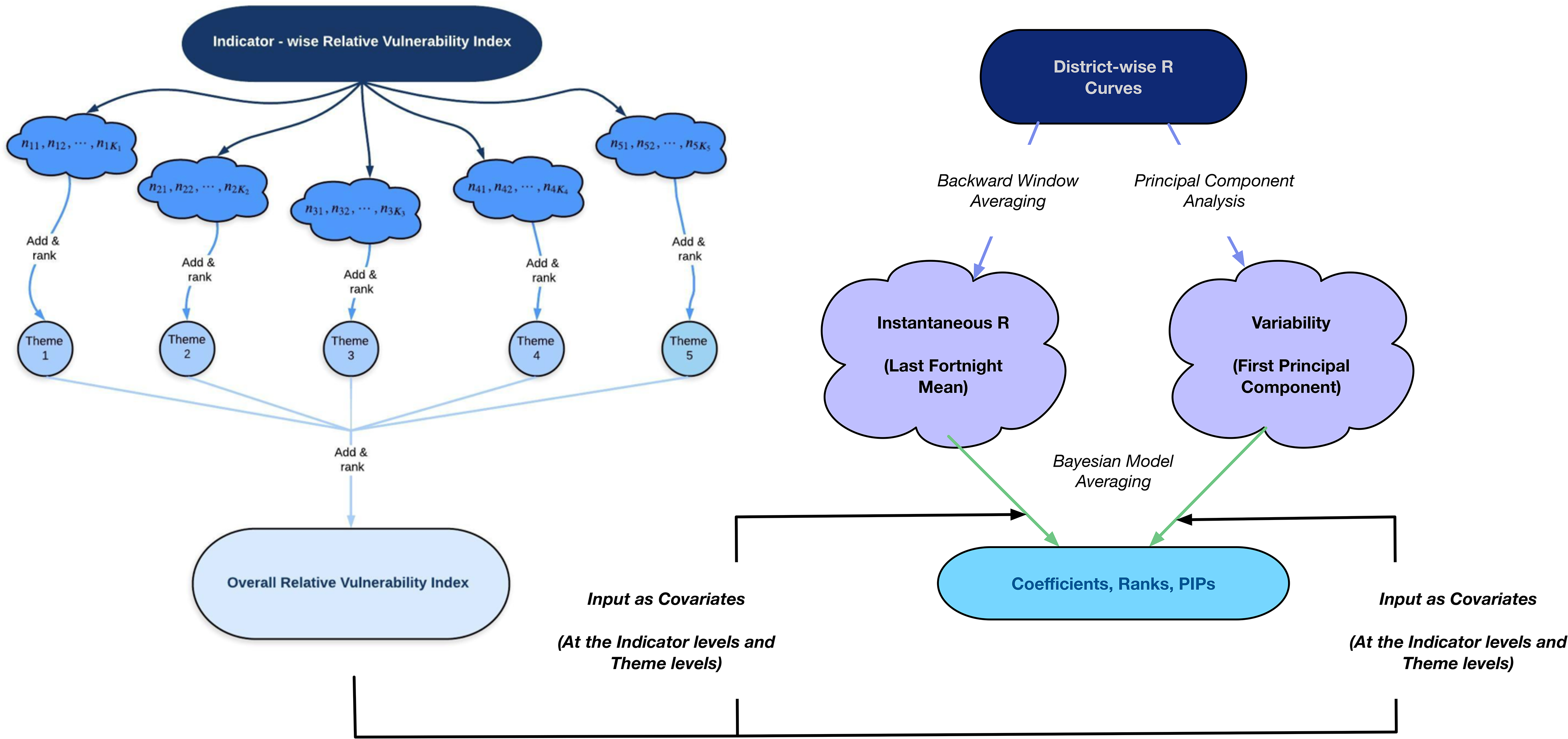
Schematic representation of data layers and approaches of the analyses performed. The left panel exhibits the three-stage procedure used to define the indicator-specific, thematic and overall Vulnerability Indices (cVIs). The right panel summarizes the processing of time-varying R to define instantaneous R (iR) and variability (vR) summaries and their usage in the Bayesian model averaging-based regression models with the cVIs as covariates.

### Estimation and summarization of time-varying reproductive number (R)

We refer to the effective reproduction number as ‘R’ throughout, which is similar to the concept of R_0_ in typical compartmental epidemiologic models^30^ but different in the sense that R_0_ is a constant that is inherent to the virus and the affected population and is not time-varying or impacted by interventions such as social distancing or lockdown, which is the case for R. We compute two scalar summaries of location and dispersion of the time series thus obtained, named instantaneous R (*iR*, Supplementary Text Equation 4) and variability in R^31^ (*νR*, Supplementary Text Equation 5) respectively, that are used in downstream regression models. The iR captures the average behaviour of the R-curve over the past two weeks from the last observed time point.

### Regression analyses using summaries of time-varying R profiles

We undertake regression analyses using the summaries of R profiles (iR and vR) as response variables and the cVIs (theme-specific and within-theme) as our covariates, to identify the associations of these cVIs with the R summaries and estimate the extent to which changes in the cVIs impact them. The cVIs corresponding to the themes socioeconomic and demographic indices (cVI_SD_), housing and hygiene conditions (cVI_HH_), preparedness for COVID (cVI_PC_), epidemiological factors (cVI_EF_), and availability of health care (cVI_AH_) lead to a final regression model of the following form,

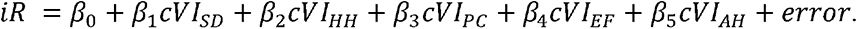

We used a Bayesian procedure to fit our regression models ^32^ and details regarding the prior structures and model fitting procedure are in Supplementary Text Equation 6. Briefly, the *β*’s quantify the amount of change in iR due to one unit change in the corresponding vulnerability index (i.e. movement from least vulnerable to most vulnerable). We used posterior inclusion probabilities (PIPs) as estimates of variable importance in the fitted model, summarizing the relative contributions of the covariates towards explaining the variability in iR across districts. We also fit similar models using iR as the response variable and within-theme cVIs as the covariates (five models, one for each theme). The details for these models are in Supplementary Text Equation 7. Correspondingly, we fit overall (Supplementary Text Equation 8) and theme-specific (Supplementary Text Equation 9) models using vR as the response.

### Ethics and patient/public involvement

Our study does not involve the participation of patients or any members of the public. All data used in this study are aggregated and publicly available, as listed in the beginning of this section and in Supplementary Table 1.

## RESULTS

### Interpreting COVID-19 susceptibility and vulnerability indices

Using the indicators listed in Figure 1 and Supplementary Table 1, we developed an area-based composite COVID-19 susceptibility and vulnerability indices at the district level-for Odisha, with a framework towards providing policy makers with some indication on which districts are likely to be most susceptible or vulnerable to a COVID-19 outbreak and specifically where the Government should potentially target its resources and accordingly plan data-driven intervention strategies. The elicited results from these indices are presented below. We first summarize the case incidence data across the 30 districts of Odisha at both state and district levels between the dates May 1, 2020 – Apr 15, 2021 in Figure 3. Figure 3A indicates that there is a spike in cases in the month of May following which during the last phase of lockdown and the first phase of unlock till June end, the reported cases remained relatively stable. This could be due to either strict lockdown and associated measures or undercounting of cases due to limited testing capacity. Following Unlock 2.0 in the month of July there is a steady rise in the cases reported. From the end of July, through Unlock 3.0 till early September, a steep rise in the reported cases is witnessed. Figure 3B shows that from mid-September onwards, the rate of cases being reported has apparently slowed down again. In more recent times, a new spike is being observed, beginning during April 2021. Figure 3C indicates that during lockdown, the SARS-CoV-2 was mostly active in districts such as Cuttack, Deogarh, Dhenkanal, Kalahandi, Koraput, and Puri (showing high values of R in red). They continue to show high values of R in the subsequent unlock phases as well, excluding Koraput, which shows days with relatively low values of R in the period after Unlock 4.0 indicating that the district has managed to reduce the incidence. All districts showed moderate and controlled values of R during Unlock 5.0 to 7.0, but the R tended to increase in the initial months of 2021. Figure 3D indicates that as of the first fortnight of April 2021, all the districts experience > 1, i.e., further growth of the pandemic over this period.

**Figure 3:**
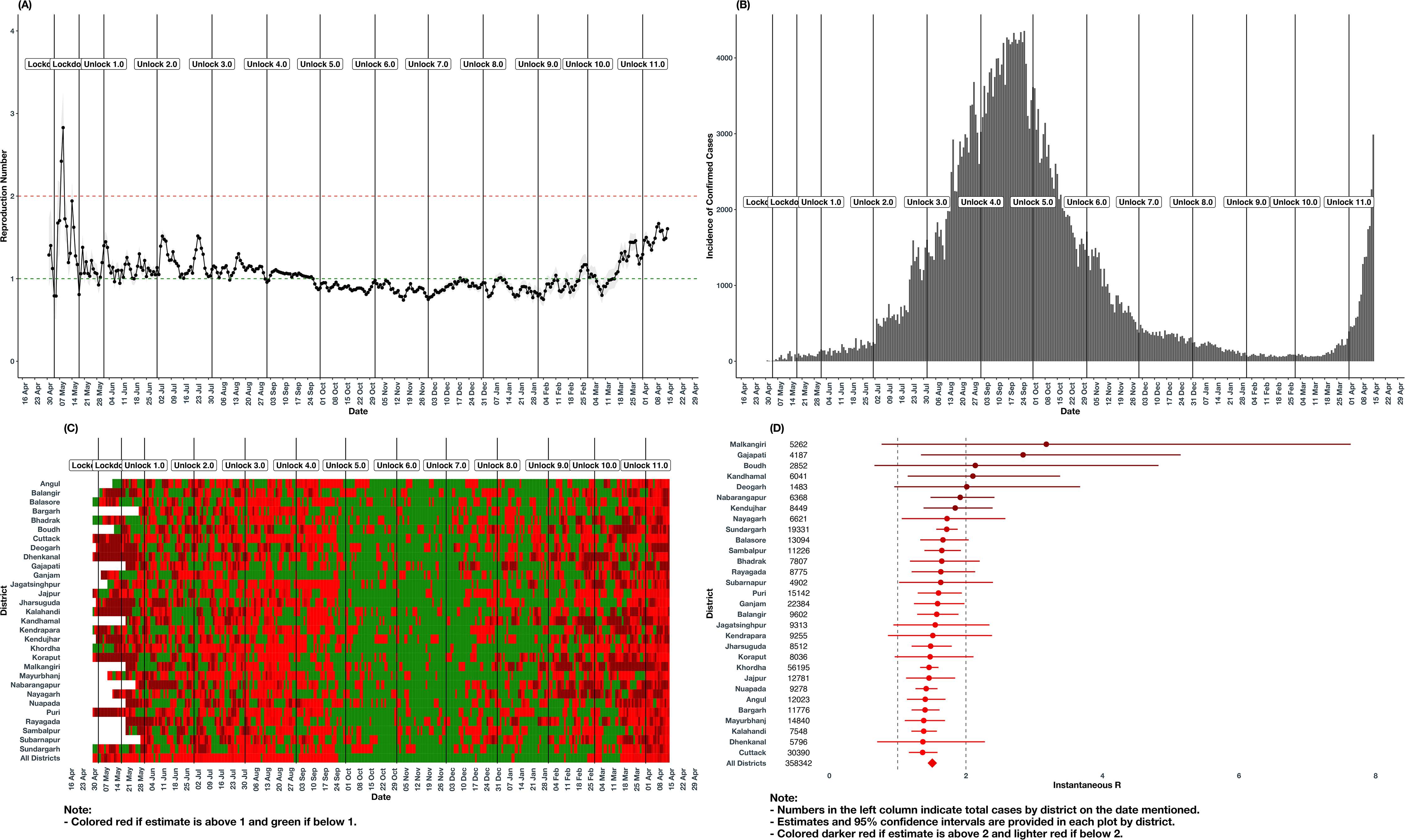
Summary of estimated time-varying R and COVID-19 incidence across Odisha districts. **Panel A.** Estimated time-varying R and 95% CI during May 1, 2020 – April 15, 2021 for Odisha. The red and the green horizontal lines indicate R = 2 and R = 1, respectively. The vertical lines indicate the beginnings of the national lockdown and unlock periods as labelled. **Panel B.** Daily incidence of reported COVID-19 cases during May 1, 2020 – April 15, 2021 for Odisha. The vertical lines indicate the beginnings of the national lockdown and unlock periods as labelled. **Panel C.** The heat map of discretized estimated time-varying R shows the progression of COVID-19 in the state of Odisha over time. The map is read from left to right, and colour coded to show the relative numbers of new cases from May 1, 2020 – April 15, 2021 by the 30 districts of the state. **Panel D.** The forest plot of instantaneous R and 95% CI at April 15, 2021 for the 30 districts and the entire state of Odisha. The left and right vertical lines indicate R = 1 and R = 2, respectively.

Figure 4 shows that the spatial map of the of the cVI’s along with the epidemic spread across all the districts of the state, with the colour grading of the districts indicating the oVI values and the sizes of the dots indicating iR values. Clearly, all the districts have iR larger than 1. On observation of the spatial plots for overall vulnerability for Odisha, some dissimilarities among the vulnerability index and district wise cases are witnessed. The districts of Malkangiri, Nabrangapur, Rayagada, Mayurbhanj have relatively high oVI values, as seen in Figure 4A. Within these, Mayurbhanj shows a relatively low iR value. On the other hand, districts like Sambalpur and Balasore have high values of iR despite having low oVI values. Not all these results are along the direction of our expectations. Hence, we further explore the individual theme level indices based on Figure 4B-F, with the colours of the districts now indicating the theme-specific cVI values.

**Figure 4:**
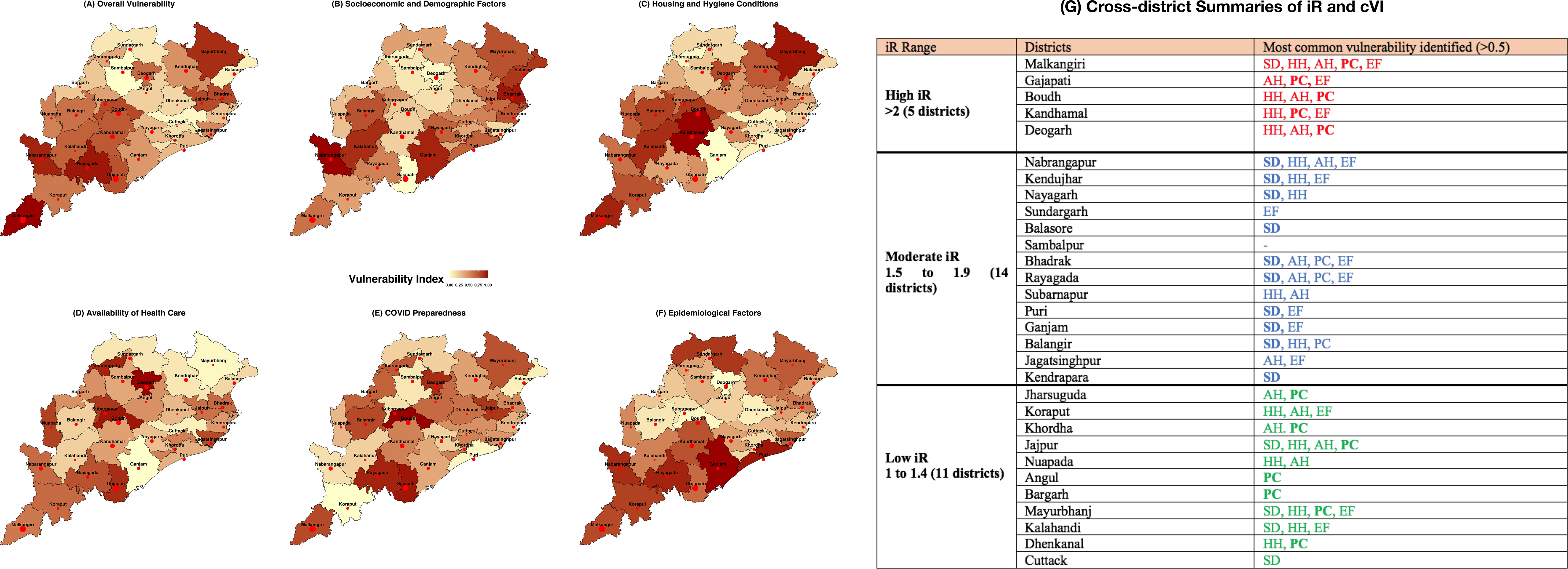
Summary of overall and themed vulnerability indices and iR across the 30 districts of Odisha. Panel A corresponds to the overall vulnerability index and Panels B-F depict the five themed vulnerability indices, as mentioned in the individual panel titles. The size of the red dots are proportional to the corresponding instantaneous R (iR) at April 15, 2021. Panel G summarizes the clusters of districts colour-coded and categorized by iR with associated vulnerability themes - socioeconomic and demographic (SD), housing and hygiene conditions (HH), preparedness for COVID (PC), epidemiological factors (EF), and availability of health care (AH). Colours red, blue, and green indicate high, moderate, and low iR districts respectively. The theme in bold is the most commonly occurring vulnerability in the cluster.

For cVI_SD_, districts like Ganjam, Balangir, and Kendrapara show relatively high cVI with relatively low values of iR compared to the five districts with highest iR. Districts such as Gajapati, Sambalpur, Kandhamal, and Boudh show high values of iR despite having low cVI values for this theme (Figure 4B). For cVI_HH_, districts like Malkangiri, Kandhamal, Boudh have high cVI values as well as high values of iR. On the other hand, districts like Puri, Cuttack and Bargarh have low iR values as well low values of cVI (Figure 4C). For cVI_AH_, we see that the expected pattern of relatively high cVI associated with high iR is generally followed. The districts of Gajapati, Deogarh, and Boudh fall in this class. However, districts such as Kendujhar and Nuapada are exceptions (Figure 4D). Similarly, for cVI_PC_, districts like Gajapati, Deogarh and Malkangiri have high values of both cVI and IR. On the other hand, districts like Koraput, Puri, and Subarnapur have low values of both metrics. However, there are exceptions where the iR and cVI are in opposite directions such as the districts of Balasore, Sambalpur, and Sundargarh (low cVI, high iR) and Rayagada and Jajpur (high cVI, low iR) (Figure 4E). For cVI_EF_, we see that the districts of Ganjam, Puri, Rayagada, and Koraput show relatively low iR despite a high cVI. Boudh, Sambalpur, and Balasore, on the other hand, show low cVI with high iR values (Figure 4F).

All districts have an iR greater than 1 in the second wave. Figure 4G shows that three clusters of districts with most common vulnerability were identified where red, blue, and green colour indicates high, moderate, and low iR districts respectively. The theme in bold is the most commonly occurring vulnerability in the cluster. Districts with relatively higher iR are more influenced by preparedness of COVID, housing and hygiene and availability of healthcare. In other districts with moderate iR, socio-demographic and epidemiological factors have more influence. Districts with relatively lower iR are more influenced by preparedness for COVID. District preparedness indicators influence R more and this is further accentuated by socio demographic and house hold variables.

We also observed clustering patterns in the overall as well as individual theme spatial plots. Similar shades of VI values are clustered in regions. Highly vulnerable districts (darker shades) are clustered around the southern and south-western regions of the state whereas lower vulnerability districts (lighter shades) are clustered near the central and northern part of state in case of overall vulnerability. Similar patterns are exhibited by theme level vulnerability indices like epidemiological factors, socioeconomic and demographic factors. COVID preparedness and housing and hygiene condition themes display different clustering patterns. These two sets of clusters exhibit differing sets of iR values. Nevertheless, the analysis devised from Figure 4 is largely correlative, and the interpretations are based on visual inspections. Hence, in order to understand the explicit dependence between the covariates (cVIs) and responses (iR and vR), we present the results of our Bayesian linear regression analysis next.

Figure 5 and Supplementary Figure 1 summarize the relative ranking for the overall and theme-specific vulnerability indices in terms of their PIPs along with the signs of the estimated regression coefficients with iR as the response. In the absence of any other information, since the cVIs are all interpretable in ‘the higher the worse’ direction, we would largely expect the signs to be positive. Figure 5A displaying the relative importance of the different theme-specific cVIs confirms that the cVI_HH_ is the ranked the highest, followed closely by cVI_PC_ and cVI_EF_. Only one out of these five themes cVIs yield a negative coefficient – cVI_SD_ (*β* = −0.04, *sd* = 0.15)

**Figure 5:**
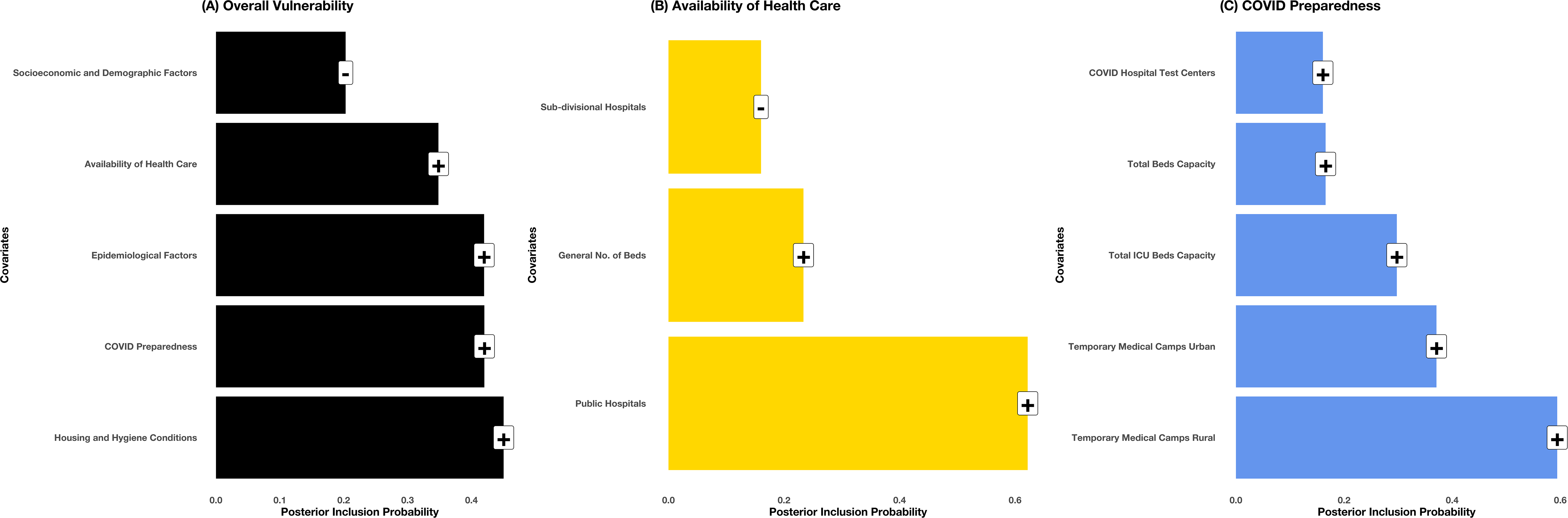
Summary of regression models for instantaneous R with the vulnerability indices as covariates. Panel A corresponds to the themed vulnerability indices constituting the overall vulnerability index and Panels B-C depict the indicators within the themes ‘Availability of Health Care’ and ‘COVID Preparedness’, as mentioned in the individual panel titles. In each case, a Bayesian model averaging-based linear regression model is fit using instantaneous R as response and the indicators/themes as covariates. The heights of the bars indicate the posterior inclusion probabilities for the covariates in the fitted models, and the labels on top of the bars indicate the signs of the estimated coefficient as obtained in those models.

To further investigate the actual association of the themed cVIs on iR, we also summarize the theme-specific models in Figure 5B-C and Supplementary Figure 1A-C. Except for cVI_SD_ and cVI_EF_, most of the indicators in all the other groups show intuitive (positive) direction of the coefficients. For each theme, a few indicators stand out, such as – literacy rate (cVI_SD_), clean fuel and improved drinking (cVI_HH_), and public hospitals (cVI_AH_). Specifically, for Figure 5C, all cVI_PC_ are calculated in a way such that the higher the cVI, the lower the availability of the corresponding resource is for a district, and vice-versa. As can be seen from the signs of the estimated regression coefficients on top of the bars in Figure 5C, for each of these cVIs, an increment in vulnerability (decrease in availability of the resource) causes the iR, and hence the pandemic spread, to increase. For example, for the cVI with the highest importance in this category, the number of temporary medical camps set up in rural regions, a unit increment in the cVI (movement from least vulnerable to most vulnerable among all the districts) increases the mean of the iR by an estimated quantity of 0.33. Supplementary Figure 2 offers the summaries of the similar regression models with vR as the response.

## DISCUSSION

The present study makes a novel attempt to examine heterogeneities in the underlying socioeconomic vulnerabilities related to COVID-19 risk across districts in the eastern state of Odisha using a composite index. Using districts in Odisha as an example, we have developed a cVI that can be adopted by the healthcare, public health and data science communities for designing more effective intervention strategies. Mapped to the districts, the vulnerability indices provide information that is useful for emergency response planning and mitigation at a relatively granular level and can help support response planning for the current and future epidemics (Figure 4G).

The heat map (Figure 3C) shows if the districts show patterns of “red” or “green” zones, indicating the burden of cases over the pandemic timeline. From the end of September 2020 cases begin to decline gradually plateauing towards the end of December 2020. From April 2021, all districts show a high burden of cases as well as high values of iR. In the beginning of April 2021, increased infection rates were reported from the western and southern border districts.^33^ The pattern of vulnerability identified show that the majority of the districts that are tribal^34^ as well as vulnerable to natural disasters^35 36^ also show high overall and socio-economic vulnerability and high number of cases for COVID-19. Many tribal districts display vulnerability across all themes. Across the state higher vulnerability districts are mostly clustered in the southern and south-western region of the state. Our findings about clustering of districts has been reflected in previous studies which have identified clusters of vulnerable districts to COVID-19 at the national level in India^37^ as well as social and epidemiologically vulnerable clusters at the sub-county level in Kenya.^7^ Another study at the subnational level also identified clusters of socially vulnerable districts to the Ebola virus disease (EVD) in Africa.^38^

### Socioeconomic and demographic composition

Poor socioeconomic conditions may be associated with a poorer recovery, reflecting the importance of this theme. In addition to such indicators, access to impaired health-care services and facilities can also significantly deteriorate immune response and patient recovery. Most districts that report a poor literacy rate as compared to the state average also have a higher iR.^39^

### Housing and hygiene conditions

The lack of proper home structure and the lack of access to minimum resources, such as water and basic sanitation in districts can increase the risk of illness due to COVID-19, as observed with other respiratory diseases.^40^ For example, poor quality housing is associated with certain health outcomes, damp housing can lead to respiratory diseases such as asthma while overcrowding can result in higher infection rates.^41^ Recent research has identified SARS-CoV-2 in the faeces of COVID-19 patients suggesting possible ‘faecal□mucosal transmission’ in public toilets or areas with poor sanitation. Most districts which show high number of cases in the second wave have poor coverage of sanitation (Malkangiri-17%, Nabrangapur-16%, Kandhamal-17%, Boudh-16%) as compared to those districts reporting lower cases (Ganjam-41%, Cuttack-39%, Khordha-47%).^19 42^ Further, previous research has demonstrated that lack of access to clean water and sanitation facilities leads to exacerbation of viral transmission of viral infections like the respiratory Avian influenza H7N9 virus.^43^ As per the National Family Health Survey (NFHS-4) in 2016, Odisha has one of the of the highest rates (65%) for open defecation. There is a significant urban–rural disparity with the proportion practising open defecation ranging from 28.3% in urban to 72.4% in rural areas in Odisha.^19^ Many of the vulnerable districts have large populations living in rural areas where open defecation is common. It is important to generate public awareness about the risk of COVID-19 and precautions to avoid the same. This is further supported by the findings related to safe sanitation practices and exposure to mass media that seem to be associated with significantly lower risk of COVID-19 in other parts of India.

### Availability of health-care facilities

Access to healthcare is lower in disadvantaged and marginalised communities - even in countries which have universal healthcare systems.^44^ In Brazil, unequal distribution in resources have resulted in regional and social healthcare inequalities.^45^ The Ebola outbreak in Africa (2014-2016) has demonstrated the importance of timely and quality healthcare from resilient health systems to prevent, detect, and respond to diseases of pandemic potential.^46^ The state of Odisha has 83% population living in rural areas and has a deficit of health infrastructure. The state has a significant shortfall of doctors, specialists and health human resources in rural areas with one Sub-centre (SC) for 5,551 people, one Primary Health Centre (PHC) for 28,822 persons and one Community Health Centre (CHC) covering a population of 98,469.^21^ This deficit is more pronounced in the Aspirational Districts (ADs) which have limited coping-up capacities.^8^ Our model shows that in the second wave certain aspirational districts have recorded a greater number of cases. However, our findings reveal that infrastructure measures taken for COVID-19 preparedness are not significantly associated with instantaneous R. Preparedness of COVID was an important factor in the 11 districts that have reported lower iR (Figure 4G). The Government of Odisha adopted a decentralised approach by setting up temporary medical camps (TMC) for the migrant influx at the Gram Panchayat level to arrest community spread of the virus. Such camps enable early detection and action to contain the spread of infection. Historically, camps offering institutional quarantine facilities have been at the core of multicomponent strategies for controlling communicable disease outbreaks.^47^

### Epidemiological factors

In the COVID-19 pandemic, obesity was identified as a major risk factor has been shown associated with disease progression.^48^ Research has shown that people with underlying uncontrolled medical conditions such as diabetes; hypertension; lung, liver, and kidney disease; cancer patients on chemotherapy; smokers; transplant recipients; and patients taking steroids chronically were at increased risk of COVID-19 infection.^49^ Brazil reported a prevalence of 83% of comorbidities in around 17,752 COVID related deaths, with the leading two comorbidities being chronic heart disease and diabetes.^50^ Odisha also had a prevalence of 33.7% of obesity rates.^19^ Districts such as Deogarh, Boudh, and Nabrangapur that showed high number of cases in both the waves also have significant numbers of people living with co-morbidities such as obesity, diabetes, and hypertension.^19^ While developing responses to tackle the current epidemic and future pandemics it would be useful to note that districts that have high epidemiological susceptibility need enhanced vigilance (through special rapid-action teams) to avoid transmissions..

## CONCLUSION

This paper constructs a multidimensional vulnerability index to account for both long-term structural vulnerabilities as well as the recent weaknesses uncovered by the pandemic. It contributes to the debate on vulnerability measurement by contrasting a narrow focus on epidemiological or environmental vulnerability with a multi-dimensional approach to assessing vulnerabilities at granular sub-regional levels. The vulnerability index has highlighted the relative vulnerability of certain districts as compared to others. Acknowledging the diversity, varying needs and priorities of the different districts in this state, our study provides information that can help detailed planning at subnational levels. Vulnerability assessment such as the cVI’s proposed in this paper, can provide an understanding of real-world situations and be used for pandemic planning at a subnational level for other states as well as in other LMICs.

### Limitations

Data-related limitations were encountered when computing the vulnerability indices due to use of secondary data that are subject to constant variation. Several variables including mobility between districts could not be accessed during the analysis. Also, the most recent data from the national surveys are not yet available and might not have captured vulnerability well in certain districts where rapid changes may have occurred. Further, due to the lack of granular data, the epidemiological indices data are not updated to 2021 and their trends are likely to have changed. Along with the known issue of underreporting of COVID-19 cases due to the limited availability of tests and the capacity of local surveillance services, there were deficiencies in investigating deaths due to the disease, with possible underreporting there as well. Therefore, a similar analysis performed on a bigger and possibly more representative sample obtained via large-scale population-level testing may have yielded better explanations. Fortunately, our analysis pipeline offers a dynamic Bayesian option to update the inferences with incoming data providing more granular, detailed, and in-depth views of the true scenario, and can be adapted suitably to any other geographical region of interest in context of the disease.

## Data Availability

All our source codes and processed data have been made freely available to the broader research community at.

https://github.com/bayesrx/COVID_vulnerability_India.

## AUTHOR CONTRIBUTIONS

RP and VB conceptualized the project. RP conceptualised the idea of the index, lead the writing of the introduction discussion and conclusion. VB conceptualised the analyses pipeline with RB leading the analyses and writing the results along to with AB, SB, SM and AA. SL helped in writing of the discussion section. KS helped in accessing and cleaning data used for constituting the index. SR reviewed the manuscript and provided important inputs on all sections of the paper.

## ACKNOWLEDGEMENTS

We would like to acknowledge all the sources that made the data used for our analyses available, including: COVID-19 India Dashboard, National Family Health Survey 2015–16, Census of India 2011, Rural Health Statistics 2018, Health management information system (HMIS) 19-20, Economic Survey of Odisha, Directorate of Economics & Statistics, Odisha, India 11-12, Directorate of Health Services, Department of Health and Family Welfare, Government of ODISHA website, and COVID Dashboard Govt. Of Odisha.

## LICENCE STATEMENT

I, the Submitting Author has the right to grant and does grant on behalf of all authors of the Work (as defined in the below author licence), an exclusive licence and/or a non-exclusive licence for contributions from authors who are: i) UK Crown employees; ii) where BMJ has agreed a CC-BY licence shall apply, and/or iii) in accordance with the terms applicable for US Federal Government officers or employees acting as part of their official duties; on a worldwide, perpetual, irrevocable, royalty-free basis to BMJ Publishing Group Ltd (“BMJ”) its licensees and where the relevant Journal is co-owned by BMJ to the co-owners of the Journal, to publish the Work in BMJ Global Health and any other BMJ products and to exploit all rights, as set out in our licence.

The Submitting Author accepts and understands that any supply made under these terms is made by BMJ to the Submitting Author unless you are acting as an employee on behalf of your employer or a postgraduate student of an affiliated institution which is paying any applicable article publishing charge (“APC”) for Open Access articles. Where the Submitting Author wishes to make the Work available on an Open Access basis (and intends to pay the relevant APC), the terms of reuse of such Open Access shall be governed by a Creative Commons licence – details of these licences and which Creative Commons licence will apply to this Work are set out in our licence referred to above.

## REFERENCES

1. Zhang T, Wu Q, Zhang Z. Probable Pangolin Origin of SARS-CoV-2 Associated with the COVID-19 Outbreak. Current biology : CB 2020;30(8):1578–78. doi: 10.1016/j.cub.2020.03.063

2. COVID-19 India. Coronavirus Outbreak in India - covid19india.org 2021 [Available from: https://www.covid19india.org/ accessed 8 February 2021.

3. The World Bank. Population density (people per sq. km of land area) 2021 [Available from: https://data.worldbank.org/indicator/EN.POP.DNST accessed 26 May 2021.

4. Pal R, Yadav U. COVID-19 Pandemic in India: Present Scenario and a Steep Climb Ahead. Journal of primary care & community health 2020;11:2150132720939402–02. doi: 10.1177/2150132720939402

5. Ebrahimi M, Malehi AS, Rahim F. COVID-19 Patients: A Systematic Review and Meta-Analysis of Laboratory Findings, Comorbidities, and Clinical Outcomes Comparing Medical Staff versus the General Population. Osong public health and research perspectives 2020;11(5):269–79. doi: 10.24171/j.phrp.2020.11.5.02

6. Lewnard JA, Liu VX, Jackson ML, et al. Incidence, clinical outcomes, and transmission dynamics of severe coronavirus disease 2019 in California and Washington: prospective cohort study. BMJ (Clinical research ed) 2020;369:m1923–m23. doi: 10.1136/bmj.m1923

7. Macharia PM, Joseph NK, Okiro EA. A vulnerability index for COVID-19: spatial analysis at the subnational level in Kenya. BMJ global health 2020;5(8):e003014. doi: 10.1136/bmjgh-2020-003014

8. Imdad K, Sahana M, Rana MJ, et al. A district-level susceptibility and vulnerability assessment of the COVID-19 pandemic’s footprint in India. Spatial and spatio-temporal epidemiology 2021;36:100390–90. doi: 10.1016/j.sste.2020.100390 [published Online First: 2020/11/08]

9. Prashanth NS, Marchal B, Kegels G, et al. Evaluation of capacity-building program of district health managers in India: a contextualized theoretical framework. Frontiers in public health 2014;2:89–89. doi: 10.3389/fpubh.2014.00089

10. Acharya R, Porwal A. A vulnerability index for the management of and response to the COVID-19 epidemic in India: an ecological study. The Lancet Global health 2020;8(9):e1142–e51. doi: 10.1016/S2214-109X(20)30300-4 [published Online First: 2020/07/16]

11. The L. Redefining vulnerability in the era of COVID-19. Lancet (London, England) 2020;395(10230):1089–89. doi: 10.1016/S0140-6736(20)30757-1

12. Kamath S, Kamath R, Salins P. COVID-19 pandemic in India: challenges and silver linings. Postgraduate Medical Journal 2020;96(1137):422-23. doi: 10.1136/postgradmedj-2020-137780

13. Wilkinson A, Ali H, Bedford J, et al. Local response in health emergencies: key considerations for addressing the COVID-19 pandemic in informal urban settlements. Environment and Urbanization 2020;32(2):503–22. doi: 10.1177/0956247820922843

14. Banik R, Rahman M, Sikder T, et al. COVID-19 in Bangladesh: public awareness and insufficient health facilities remain key challenges. Public health 2020;183:50–51. doi: 10.1016/j.puhe.2020.04.037 [published Online First: 2020/05/07]

15. Buheji M, da Costa Cunha K, Beka G, et al. The Extent of COVID-19 Pandemic Socio-Economic Impact on Global Poverty. A Global Integrative Multidisciplinary Review. American Journal of Economics 2020;10(4):213–24. doi: 10.5923/j.economics.20201004.02

16. Precision for COVID. COVID-19 Community Vulnerability Index (CCVI) 2021 [Available from: https://precisionforcovid.org/ccvi accessed May 26 2021.

17. Census of India Website : Office of the Registrar General & Census Commissioner, India. Censusindia.gov.in. 2011 [Available from: https://censusindia.gov.in/2011-common/censusdata2011.html accessed May 26 2021.

18. Economic Survey of Odisha, Directorate of Economic & Statistics, Odisha India 11-12. 2012 [Available from: http://desorissa.nic.in/pdf/lt_publication/Economic_Survey_2011_12.pdf accessed May 26 2021.

19. National Family Health Survey 2015-16 India 2016 [Available from: http://www.rchiips.org/nfhs accessed May 26 2021.

20. Rural Health Statistics. Open Government Data (OGD) Platform India. 2018 [Available from: https://data.gov.in/dataset-group-name/rural-health-statistics accessed May 26 2021.

21. HMIS-Health Management Information System. 2020 [Available from: https://hmis.nhp.gov.in/#!/ accessed May 26 2021.

22. COVID-19: Odisha State Dashboard. 2021 [Available from: https://statedashboard.odisha.gov.in/ accessed May 26 2021.

23. Directorate of Health Services, Department of Health and Family Welfare. 2021 [Available from: https://main.mohfw.gov.in/sites/default/files/Final%20RHS%202018-19_0.pdf accessed May 26 2021.

24. United Nations Development Programme. The Social and Economic Impact of Covid-19 in the Asia-Pacific Region | UNDP. 2021 [Available from: https://www.undp.org/content/undp/en/home/librarypage/crisis-prevention-and-recovery/the-social-and-economic-impact-of-covid-19-in-asia-pacific.html accessed May 26 2021.

25. Wadhera RK, Wadhera P, Gaba P, et al. Variation in COVID-19 Hospitalizations and Deaths Across New York City Boroughs. JAMA 2020;323(21):2192–95. doi: 10.1001/jama.2020.7197

26. Galobardes B, Shaw M, Lawlor DA, et al. Indicators of socioeconomic position (part 2). Journal of epidemiology and community health 2006;60(2):95–101. doi: 10.1136/jech.2004.028092

27. Rosenthal PJ, Breman JG, Djimde AA, et al. COVID-19: Shining the Light on Africa. The American journal of tropical medicine and hygiene 2020;102(6):1145–48. doi: 10.4269/ajtmh.20-0380

28. United Nations Development Programme. Human Development Data Story. 2021 [Available from: http://hdr.undp.org/sites/default/files/covid-19_and_human_development.pdf accessed May 26 2021.

29. Flanagan BE, Gregory EW, Hallisey EJ, et al. A Social Vulnerability Index for Disaster Management. Journal of Homeland Security and Emergency Management 2011;8(1) doi: 10.2202/1547-7355.1792

30. Tang L, Zhou Y, Wang L, et al. A Review of Multi-Compartment Infectious Disease Models. International statistical review = Revue internationale de statistique 2020;88(2):462–513. doi: 10.1111/insr.12402 [published Online First: 2020/08/03]

31. Pearson K. LIII. On lines and planes of closest fit to systems of points in space. The London, Edinburgh, and Dublin Philosophical Magazine and Journal of Science 1901;2(11):559–72. doi: 10.1080/14786440109462720

32. Zeugner S, Feldkircher M. Bayesian Model Averaging Employing Fixed and Flexible Priors: TheBMSPackage forR. Journal of Statistical Software 2015;68(4) doi: 10.18637/jss.v068.i04

33. Das P. As the second wave hits Odisha and new COVID-19 cases increase dramatically, the government resorts to night curfews and weekend lockdowns to contain the spread.: The Hindu Frontline; 2021 [Available from: https://frontline.thehindu.com/dispatches/as-the-second-wave-hits-odisha-and-new-covid-19-cases-increase-dramatically-the-government-resorts-to-night-curfews-and-weekend-lockdowns-to-contain-the-spread/article34365189.ece accessed May 26 2021.

34. Special Development Council. 2021 [Available from: http://www.sdcodisha.in/ accessed May 26 2021.

35. Mohapatra M, Mandal GS, Bandyopadhyay BK, et al. Classification of cyclone hazard prone districts of India. Natural Hazards 2011;63(3):1601–20. doi: 10.1007/s11069-011-9891-8

36. Bahinipati CS. Assessment of vulnerability to cyclones and floods in Odisha, India: a district-level analysis. Current Science 2014:1997–2007.

37. Sarkar A, Chouhan P. COVID-19: District level vulnerability assessment in India. Clinical epidemiology and global health 2021;9:204–15.

38. Stanturf JA, Goodrick SL, Warren Jr ML, et al. Social vulnerability and Ebola virus disease in rural Liberia. PLoS One 2015;10(9):e0137208.

39. Results of District Level Household Survey - IV 2012-13 (DLHS - IV). 2013 [Available from: https://nrhm-mis.nic.in/SitePages/DLHS-4.aspx?Paged=TRUE&p_SortBehavior=0&p_ID=2025&RootFolder=%2F%2525&PageFirstRow=361&&View=%7BF8D23EC0-C74A-41C3-B676-5B68BDE5007D%7D accessed May 26 2021.

40. Sheikh A. How You Vote Can Kill YouHealth Divides: Where You Live Can Kill You By Bambra Clare Bristol (UK) : Policy Press, 2016 256 pp., $22.00. Health Affairs 2017;36(2):380–81. doi: 10.1377/hlthaff.2016.1505

41. Filho EBdS, Silva AL, Santos AO, et al. Infecções Respiratórias de Importância Clínica: uma Revisão Sistemática. REVISTA FIMCA 2017;4(1):7–16. doi: 10.37157/fimca.v4i1.5

42. Tian Y, Rong L, Nian W, et al. Review article: gastrointestinal features in COVID-19 and the possibility of faecal transmission. Alimentary pharmacology & therapeutics 2020;51(9):843–51. doi: 10.1111/apt.15731 [published Online First: 2020/03/31]

43. Zhu Z, Liu Y, Xu L, et al. Extra-pulmonary viral shedding in H7N9 Avian Influenza patients. Journal of Clinical Virology 2015;69:30–32.

44. Todd A, Copeland A, Husband A, et al. Access all areas? An area-level analysis of accessibility to general practice and community pharmacy services in England by urbanity and social deprivation. BMJ open 2015;5(5):e007328–e28. doi: 10.1136/bmjopen-2014-007328

45. Albuquerque MVd, Viana ALdÁ, Lima LDd, et al. Desigualdades regionais na saúde: mudanças observadas no Brasil de 2000 a 2016. Ciência & Saúde Coletiva 2017;22(4):1055–64. doi: 10.1590/1413-81232017224.26862016

46. Hulland EN, Wiens KE, Shirude S, et al. Travel time to health facilities in areas of outbreak potential: maps for guiding local preparedness and response. BMC medicine 2019;17(1):232–32. doi: 10.1186/s12916-019-1459-6

47. Tognotti E. Lessons from the history of quarantine, from plague to influenza A. Emerging infectious diseases 2013;19(2):254.

48. Klang E, Kassim G, Soffer S, et al. Severe Obesity as an Independent Risk Factor for COVID-19 Mortality in Hospitalized Patients Younger than 50. Obesity (Silver Spring, Md) 2020;28(9):1595–99. doi: 10.1002/oby.22913 [published Online First: 2020/08/02]

49. Centers for Disease Control and Prevention. Symptoms of COVID-19. 2021 [Available from: https://www.cdc.gov/coronavirus/2019-ncov/symptoms-testing/symptoms.html accessed May 26 2021.

50. Pachiega J, Afonso AJDS, Sinhorin GT, et al. Chronic heart diseases as the most prevalent comorbidities among deaths by COVID-19 in Brazil. Revista do Instituto de Medicina Tropical de Sao Paulo 2020;62:e45–e45. doi: 10.1590/S1678-9946202062045

