## Supplementary Document for "Role of Multi-resolution Vulnerability Indices in COVID-19 spread: A Case Study in India"

**SUPPLEMENTARY TEXT**

**Computation of relative COVID-19 vulnerability indices**

We adapt the algorithm by Acharya and Porwal, 2020 for the calculation of the relative vulnerability indices for each district.^1^ The algorithm consists of three steps, as summarized in the left panel of Figure 2 in the main paper. In the Step 1, ranks are assigned to the values of a particular indicator for different districts of the state. The ranking is done in a manner so that the ‘riskier’ the value is (relative to the other values corresponding to the same indicator), the higher is the rank assigned, i.e., if the value of the indicator for a certain district suggest minimum risk, then it is given the least rank 1, and if the value suggests maximum risk, then it is ranked the highest. Using these ranks, the *indicator-specific relative cVI* for a district is computed (Equation 1). In Step 2 we calculate *theme-specific cVI* by aggregating the *indicator-specific vulnerability index* of the indices under that particular theme (Equation 2). In the Step 3, the *overall vulnerability index (oVI)* is calculated by aggregating the *theme-specific cVI’s* (Equation 3).

Equation 1

For each of the $n$ districts of interest, and for a particular indicator denoted by$I_{j}$, let its values for the $n$ districts be$\left\{ x_{1j},x_{2j},\ldots,x_{nj} \right\}$. Now, each of these $x_{ij}$ values are assigned ranks $R_{iI_{j}}$ in such a way so that the ‘riskier’ the value is (relative to the other values corresponding to the same indicator), the higher is the value of$R_{iI_{j}}$. Thus, for the set of $n$ values of the indicator across the $n$ districts, let the ranks assigned be$\left\{ R_{1I_{j}},R_{2I_{j}},\ldots,R_{nI_{j}} \right\}$. Now, using these indicator-specific ranks for$I_{j}$, we define the *indicator-specific relative vulnerability index* for a district $D_{i}$ as:

${cVI}_{iI_{j}}=\frac{R_{iI_{j}}-1}{n-1}$.

The definition of ${cVI}_{iI_{j}}$implies that it takes a value between 0 and 1. As mentioned previously, the ranks have been assigned in such a way so that higher the value of${cVI}_{iI_{j}}$,${cVI}_{ij}$, the more vulnerable that district is w.r.t to $I_{j}$ relative to other districts; i.e., a value of ${cVI}_{iI_{j}}=1$ makes that district the most vulnerable one and a value of 0 suggests that the district is least vulnerable relative to other districts. In case of ties, the minimum value of $R_{iI_{j}}$ among the ties is assigned to the districts having the tied values.

Equation 2

For a particular theme $T_{j}$, let the indicators associated with it be$\left\{ I_{n_{j_{1}}},I_{n_{j_{2}}}\ldots,I_{n_{j_{k_{j}}}} \right\}$. After calculating the indicator-wise relative vulnerability index for the indicators grouped under $T_{j}$ (using Equation 1), we now add up those relative vulnerability index values $\left\{ {cVI}_{{iI}_{n_{j_{1}}}},{cVI}_{iI_{n_{j_{2}}},}\ldots,{cVI}_{iI_{n_{j_{k_{j}}}}} \right\}$ for a particular district $D_{i}$ and define new variables $t_{ij}$ as:

$t_{ij}=\sum_{m=1}^{k_{j}} {cVI}_{iI_{n_{j_{m}}}}$.

Thus, for theme $T_{j}$, we have values$\{t_{1j},t_{2j},\ldots,t_{nj}\}$. Now we assign ranks $R_{iT_{j}}$ to them such that the higher the value of $t_{ij},$ higher is the rank assigned and vice-versa. Based on these, we then define the *theme-wise relative vulnerability index* for theme $T_{j}$ and district $D_{i}$ as:

${cVI}_{iT_{j}}=\frac{R_{iT_{j}}-1}{n-1}$.

Here, a similar interpretation is possible as in using the same definition used for ${cVI}_{iI_{j}}$with $I_{j}$ replaced by $T_{j}$.

Equation 3

For $M$ different themes, we add up the individual indices. That is, once all the *theme-wise relative vulnerability indices* $\left\{ {cVI}_{iT_{1}},{cVI}_{iT_{2}},\ldots,{cVI}_{iT_{M}} \right\}$ are available, we aggregate the theme-wise indices for a particular district $D_{i}$ and define new variables $o_{ij}$ as:

$o_{ij}=\sum_{m=1}^{M} {cVI}_{iT_{m}}$.

Here, we assign and then use these values for obtaining increasing ranks $R_{ij}$. The same process is then applied to $o_{ij}$ to compute the *overall relative vulnerability index* ${oVI}_{ij}$ as the final aggregate across the theme-specific indices:

${oVI}_{ij}=\frac{R_{ij}-1}{n-1}$.

Here, we can also interpret ${oVI}_{ij}$ like ${cVI}_{iI_{j}}$or ${cVI}_{iT_{j}}$.

**Estimation and summarization of time-varying** re**productive number (R)**

For the $i^{th}$ location of interest (a district, in our case) the input to the EpiEstim package is represented by $\left\{ I^{i}\left( t \right), t\in\left\{ 1,\ldots,T \right\} \right\}$ where $I^{i}(t)$ indicates the incidence (count of new COVID-19 cases) in that location on day $t$, and $T$ denotes the total number of days for which this data is available. Using this input, a Bayesian estimation procedure utilizing user-supplied mean and standard deviation for the serial interval distribution yields a time series of effective reproduction number estimates $\left\{ R^{i}\left( t \right), t\in\left\{ 1,\ldots,T \right\} \right\}$. This procedure is performed for each location separately across $i\in\{1,\ldots,n\}$, say.

The details of the parameter and function choices for the estimation procedure are summarized in Supplementary Table 2. Instead of using the full time series of $\left\{ R^{i}\left( t \right), t\in\left\{ 1,\ldots,T \right\}, i\in\left\{ 1,\ldots,n \right\} \right\}$ as a response variable, we decided to compute scalar summaries of the time series at the district levels. We compute two scalar summaries, namely, a 14-day average of estimated time-varying R, called the instantaneous R (iR) (Equation 4), and the first principal component of the time-varying R, called the variability in R (vR) (Equation 5).

Equation 4

We summarize the location information the time-varying R profiles by taking a mean of the profile at a location level over a specified period of estimation and call this summary the instantaneous R ($iR$).

$iR_{K}^{i}=\frac{\sum_{t=T-K+1}^{T} R\left( t \right)}{K}, i\in\{1,\ldots,n\}$.

The normality for this metric was checked visually via density and quantile-quantile plots, as exhibited in Supplementary Figure 3.

Equation 5

Two estimated time-varying R profiles with the same computed $iR$ may represent potentially different trajectories of the pandemic depending on how dispersed the actual profile is around that representative value. Therefore, in order to obtain a summary of the variability in the R profiles, we performed a principal component analysis (PCA) on $\left\{ R^{i}\left( t \right), t\in\left\{ T-K+1,\ldots,T \right\}, i\in\left\{ 1,\ldots,n \right\} \right\}$.^2^ Denoting the matrix of these estimated R values as $P_{n\times K}, P_{it}=R^{i}\left( t \right), t\in\left\{ T-K+1,\ldots,T \right\}, i\in\left\{ 1,\ldots,n \right\}$, the PCA is performed on $P$ and the weight matrix ($W_{K\times K}$) is found as the matrix of eigenvectors of the covariance matrix of $P$. Then, we define the variability in R as the first principal component.

$vR_{K}^{i}=\left( PW \right)_{i1}, i\in\left\{ 1,\ldots,n \right\}$.

The details regarding the values of $n, K, M$ and other choices are summarized in Supplementary Table 2.

**Regression analyses using summaries of time-varying R profiles**

Our response variable is denoted by $iR_{K}^{i}, i\in\{1,\ldots,n\}$. We simplify the notations for the cVIs a bit first. For a particular theme $T_{j}$, the indicator-level covariates (cVIs) are denoted by ${cVI}_{\mathrm{iq}}^{j},i\in\left\{ 1,\ldots,n \right\},q\in\{1,\ldots,k_{j}\}$ where $k_{j}$ is the number of indicators available for that particular theme. Across the M available themes, the theme-level cVIs are denoted by ${cVI}_{ij},i\in\left\{ 1,\ldots,n \right\},j\in\{1,\ldots,M\}$. Using these, we respectively fit one overall model (Equation 6) and M theme-specific models (Equation 7) with the iR values as the response.

Equation 6

The overall model looks like the following.

$iR_{K}^{i}=\beta_{0}+\sum_{j=1}^{M} \beta_{j}{cVI}_{ij}+\epsilon_{i},i\in\left\{ 1,\ldots,n \right\}$,

$where \epsilon_{i}\sim N\left( 0,\sigma^{2} \right) iid across i, \beta_{0} and \beta_{j}s jointly follow N_{M+1}\left( \boldsymbol{0}, g\left( \sigma^{2}V^{T}V \right)^{-1} \right)$,

$i.e. the Zellner^{'}s g-prior, V being the full design matrix$,

$\log\left( \sigma\right)\sim U\left( \mathbb{R} \right), g=n$.

Equation 7

The theme-specific indicator-level model for the $j^{th}$ theme looks like the following.

$iR_{K}^{i}=\beta_{0}^{j}+\sum_{q=1}^{k_{j}} \beta_{q}^{j}{cVI}_{\mathrm{iq}}^{j}+\epsilon_{i}^{j},i\in\left\{ 1,\ldots,n \right\}$,

$where \epsilon_{i}^{j}\sim N\left( 0,\sigma_{j}^{2} \right) iid across i, \beta_{0}^{j} and \beta_{q}^{j}s jointly follow N_{k_{j}+1}\left( \boldsymbol{0}, g\left( \sigma_{j}^{2}V_{j}^{T}V_{j} \right)^{-1} \right)$,

$i.e. the Zellner^{'}s g-prior, V_{j} being the full design matrix$,

$\log\left( \sigma_{j} \right)\sim U\left( \mathbb{R} \right), g=n$.

For the vR regressions, our response variable is denoted by $vR_{K}^{i}, i\in\{1,\ldots,n\}$. Using the same notation for the cVIs as before, we respectively fit one overall model (Equation 8) and M theme-specific models (Equation 9) with the vR values as the response.

Equation 8

The overall model looks like the following.

$vR_{K}^{i}=\gamma_{0}+\sum_{j=1}^{M} \gamma_{j}{cVI}_{ij}+\delta_{i},i\in\left\{ 1,\ldots,n \right\}$,

$where \delta_{i}\sim N\left( 0,\tau^{2} \right) iid across i, \gamma_{0} and \gamma_{j}s jointly follow N_{M+1}\left( \boldsymbol{0}, g\left( \tau^{2}V^{T}V \right)^{-1} \right)$,

$i.e. the Zellner^{'}s g-prior, V being the full design matrix$,

$\log\left( \tau\right)\sim U\left( \mathbb{R} \right), g=n$.

Equation 9

The theme-specific indicator-level model for the $j^{th}$ theme looks like the following.

$vR_{K}^{i}=\gamma_{0}^{j}+\sum_{q=1}^{k_{j}} \gamma_{q}^{j}{cVI}_{\mathrm{iq}}^{j}+\delta_{i}^{j},i\in\left\{ 1,\ldots,n \right\}$,

$where \delta_{i}^{j}\sim N\left( 0,\tau_{j}^{2} \right) iid across i, \gamma_{0}^{j} and \gamma_{q}^{j}s jointly follow N_{k_{j}+1}\left( \boldsymbol{0}, g\left( \tau_{j}^{2}V_{j}^{T}V_{j} \right)^{-1} \right)$,

$i.e. the Zellner^{'}s g-prior, V_{j} being the full design matrix$,

$\log\left( \tau_{j} \right)\sim U\left( \mathbb{R} \right), g=n$.

We use the R package BMS with default choices for the bms function for fitting our models which performs a Bayesian model averaging procedure (BMA) to obtain posterior estimates, credible intervals and posterior inclusion probabilities (PIPs) for the parameters of interest.^3^ Briefly, BMA performs a search across the weighted posterior probabilities of the plausible models and selects the model with the highest posterior probability.

**SUPPLEMENTARY FIGURES**

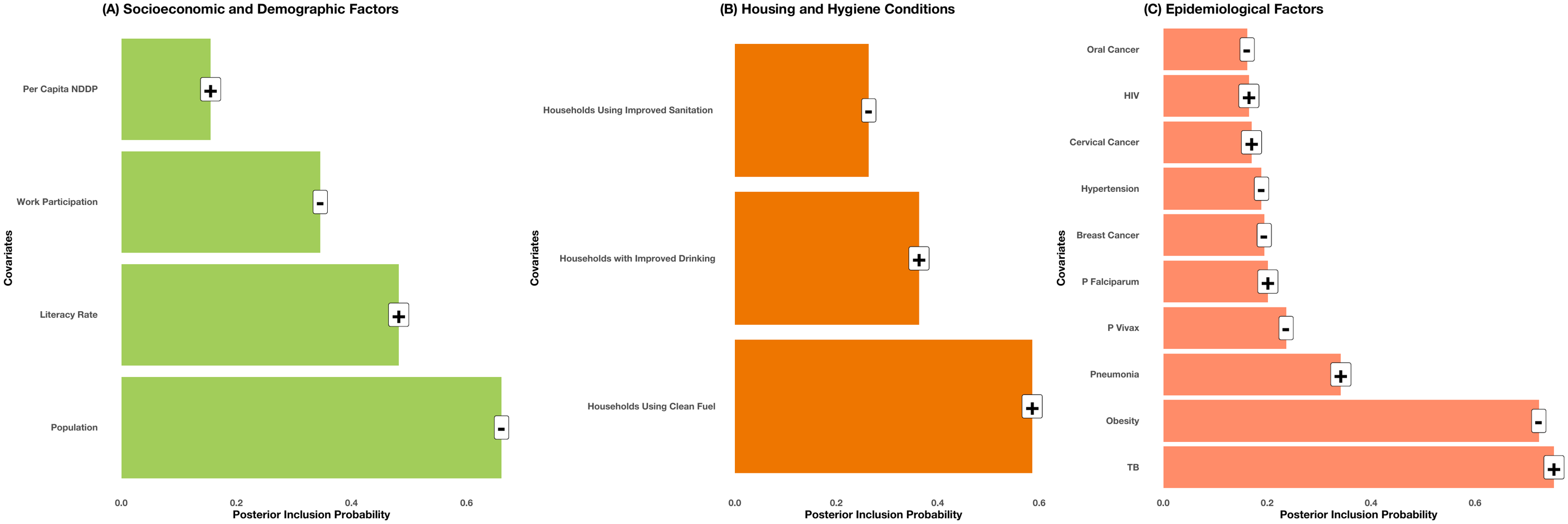

**Supplementary Figure 1: Summary of regression models for instantaneous R with the vulnerability indices as covariates.**

Panels A-C depict the indicators within the themes ‘Socioeconomic and Demographic Factors’, ‘Housing and Hygiene Conditions’ and ‘Epidemiological Factors’ respectively, as mentioned in the individual panel titles. In each case, a Bayesian model averaging-based linear regression model is fit using instantaneous R as response and the indicators/themes as covariates. The heights of the bars indicate the posterior inclusion probabilities for the covariates in the fitted models, and the labels on top of the bars indicate the signs of the estimated coefficient as obtained in those models.

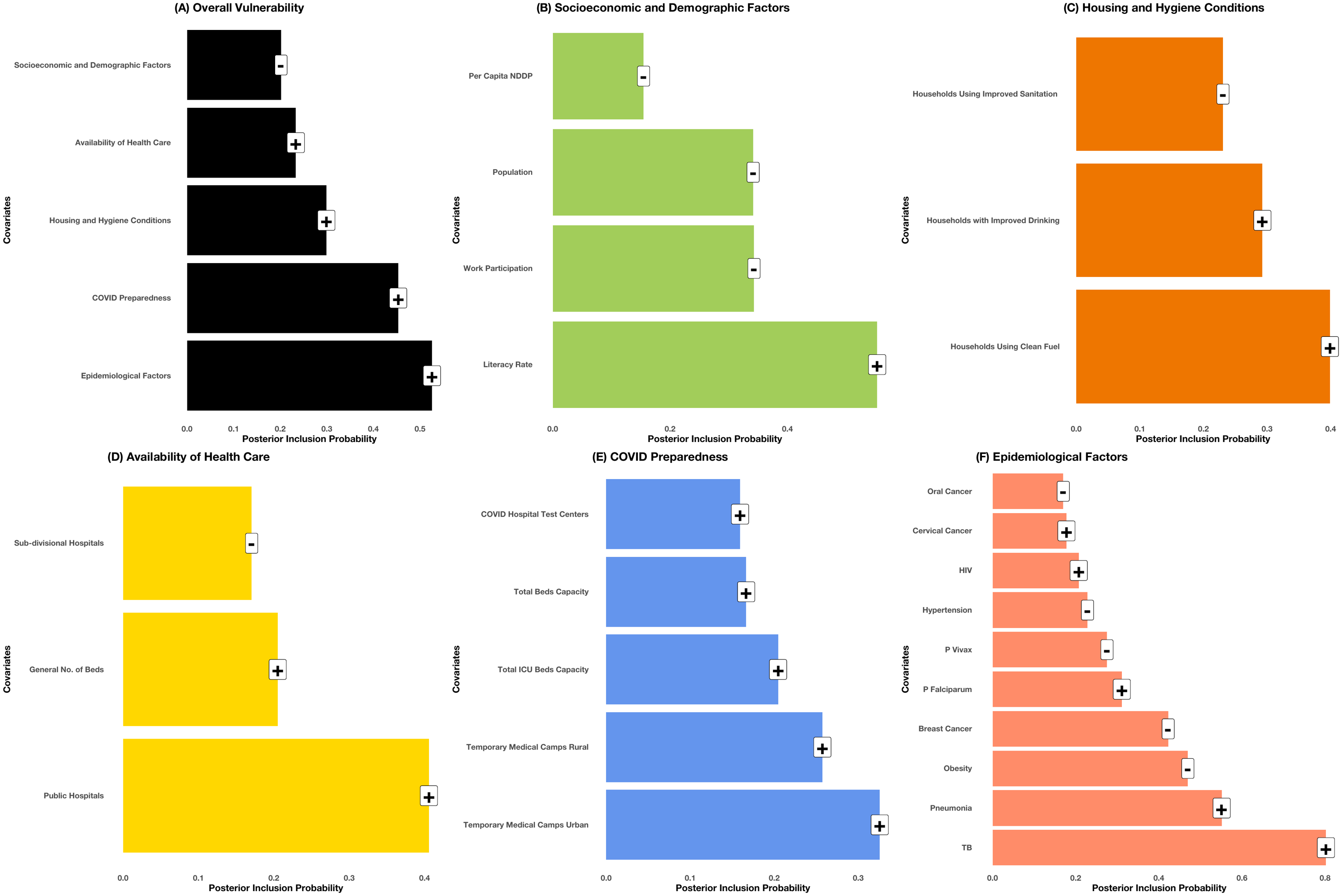

**Supplementary Figure 2: Summary of regression models for variability of R (vR) with the vulnerability indices as covariates.**

Panel A corresponds to the themed vulnerability indices constituting the overall vulnerability index and Panels B-F depict the indicators within the five themed vulnerability indices, as mentioned in the individual panel titles. In each case, a Bayesian model averaging-based linear regression model is fit using vR as response and the indicators/themes as covariates. The heights of the bars indicate the posterior inclusion probabilities for the covariates in the fitted models, and the labels on top of the bars indicate the signs of the estimated coefficient as obtained in those models.

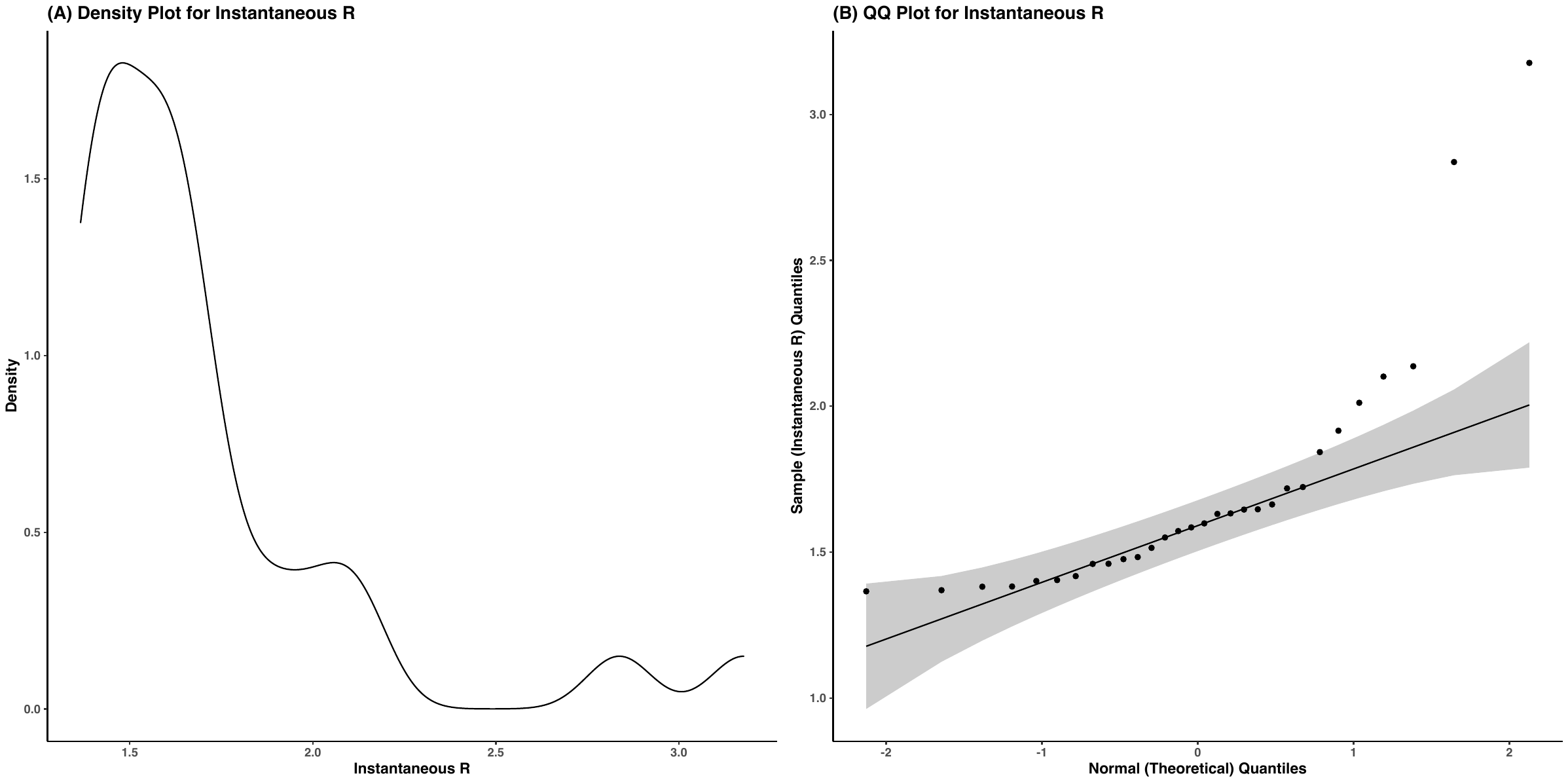

**Supplementary Figure 3: Density plot and quantile-quantile plot for the last fortnight mean of estimated R (instantaneous R or iR).**

**SUPPLEMENTARY TABLES**

**Supplementary Table 1: Summary of indicators constituting the five themed vulnerability indices along with data sources.**

| **Variable** | **Variable Description** | **Data Source** |
| --- | --- | --- |
| ***Socioeconomic and Demographic Factors (Theme 1)*** | | |
| **Population** | Calculated as a number of population (district wise) | Census of India, 2011  Population data: linearly projected population for 2019 using growth rate calculated for each district based on 2001 and 2011 census |
| **Literacy Rate** | Calculated as percentage of females who completed primary level of education | Census of India, 2011 |
| **Work Participation** | Calculated as percentage of people who are working |  |
| **Per Capita NDDP** |  | Economic Survey of Odisha, Directorate of Economic & Statistics, Odisha India 2011-12 |
| ***Housing and Hygiene Conditions (Theme 2)*** | | |
| **Households with improved Drinking Water Sources** | Calculated as percentage of households reporting improved Drinking Water Sources | National Family Health Survey-4 2015-16 |
| **Households using improved Sanitation facility** | Calculated as percentage of households reporting improved Sanitation facility |  |
| **Households using clean fuel for cooking** | Calculated as percentage of households reporting clean fuel for cooking |  |
| ***Availability of Health Care (Theme 3)*** | | |
| **Availability of public hospitals (at district level)** | Calculated as number of public hospitals (primary health center, sub-divisional and above) per 10000 population | Directorate of Health Services, Department of Health and Family Welfare, Government of ODISHA website |
| **General number of beds (at district level)** | Number of beds available per 10000 population |  |
| ***Preparedness of COVID (Theme 4)*** | | |
| **Total Beds Capacity (at district level)** | Capacity of beds per 10000 population | COVID Dashboard Govt. Of Odisha |
| **Total ICU Beds (at district level)** | ICU beds per 10000 population |  |
| **Temporary Medical Camps (at district level)** | Temporary medical camps per 10000 population |  |
| **COVID Hospital Testing Centres (at district level)** | Testing centres per 10000 population |  |
| ***Epidemiological Factors (Theme 5)*** | | |
| **Total HIV Positive** | Percentage of total HIV positive to total tested (Male + Female) | Health Management Information System (2019-20) |
| **Plasmodium Vivax Test Positive** | Percentage of plasmodium Vivax test positive to total blood smears examined |  |
| **Plasmodium Falciparum Test Positive** | Percentage of plasmodium Falciparum test positive to total blood smears examined |  |
| **Infants Deaths due to Pneumonia** | Percentage of deaths due to Pneumonia to total reported Infant deaths |  |
| **Hypertension** | Percentage of Hypertension positive to total persons examined | National Family Health Survey-4 2015-16 |
| **Cervix** | Percentage of Cervix positive to total persons examined |  |
| **Breast** | Percentage of Breast positive to total persons examined |  |
| **Oral cavity** | Percentage of Oral cavity positive to total persons examined |  |
| **Obesity** | Percentage of overweight/obese to total persons examined |  |

**Supplementary Table 2. Summary of sample size and parameter choices for the effective reproduction number estimation and subsequent regression procedures.**

| Parameter | Value Used | Justification |
| --- | --- | --- |
| *Sample Size and Related Choices* | | |
| n (Sample Size) | 30 | Data used across the 30 districts of Odisha. |
| M (Number of Themes) | 5 | Data available from across different surveys. |
| T (Number of Days) | 350 (May 1, 2020 – Apr 15, 2021) | Initial few months prior to May saw very cases in most of the Odisha districts, resulting in inflated and unstable estimates of the effective reproduction number. |
| K (iR Window) | 14 (Apr 2 – Apr 15, 2021) | K = 1, 7, 14, 30 were investigated. K = 14 yielded the most stable and close to normal distribution of the iR. |
| *Choices for estimate_R Function in Epiestim R Package^4^* | | |
| Method | parametric_SI | Known and specifiable mean and sd of the serial interval distribution, as indicated in the subsequent rows. |
| Estimation Window | 5 days | Chosen by investigating stability of the estimates and widths of the resulting confidence intervals across varying window lengths. |
| Serial Interval Prior Distribution | Gamma with mean 3.96 days and sd 4.75 days | Following recent studies on serial interval distribution of COVID-19 cases.^5^ |

**REFERENCES**

1. Acharya R, Porwal A. A vulnerability index for the management of and response to the COVID-19 epidemic in India: an ecological study. *The Lancet Global health* 2020;8(9):e1142-e51. doi: 10.1016/S2214-109X(20)30300-4 [published Online First: 2020/07/16]

2. Pearson K. LIII. On lines and planes of closest fit to systems of points in space. *The London, Edinburgh, and Dublin Philosophical Magazine and Journal of Science* 1901;2(11):559-72. doi: 10.1080/14786440109462720

3. Zeugner S, Feldkircher M. Bayesian Model Averaging Employing Fixed and Flexible Priors: TheBMSPackage forR. *Journal of Statistical Software* 2015;68(4) doi: 10.18637/jss.v068.i04

4. Cori A, Ferguson NM, Fraser C, et al. A new framework and software to estimate time-varying reproduction numbers during epidemics. *American journal of epidemiology* 2013;178(9):1505-12. doi: 10.1093/aje/kwt133 [published Online First: 2013/09/15]

5. Du Z, Xu X, Wu Y, et al. Serial interval of COVID-19 among publicly reported confirmed cases. *Emerging infectious diseases* 2020;26(6):1341.
